# Demographic and clinical profile of black patients with chronic kidney disease attending Charlotte Maxeke Johannesburg Academic Hospital (CMJAH) in Johannesburg, South Africa

**DOI:** 10.1101/2022.03.16.22272477

**Authors:** Alfred J. Meremo, Graham Paget, Raquel Duarte, Caroline Dickens, Therese Dix-Peek, Deogratius Bintabara, Saraladevi Naicker

## Abstract

**Background:** The prevalence of chronic kidney disease (CKD) is increasing worldwide; black patients have an increased risk of developing CKD and end stage kidney disease (ESKD) at significantly higher rates than other races.

**Methods:** A cross sectional study was carried out on black patients with CKD attending the kidney outpatient clinic at Charlotte Maxeke Johannesburg Academic Hospital (CMJAH) in South Africa, between September 2019 to March 2020. Demographic and clinical data were extracted from the ongoing kidney outpatient clinic records and interviews, and were filled in a questionnaire. Patients provided blood and urine for laboratory investigations as standard of care, data were descriptively and inferentially analysed using STATA version 17. Multivariable logistic regression analysis was used to identify demographic and clinical data associated with advanced CKD.

**Results:** A total of 312 black patients with CKD were enrolled during the study period; 58% patients had advanced CKD, of whom 31.5 % had grossly increased proteinuria, 96.7 % had hypertension, 38.7 % had diabetes mellitus and 38.1 % had both hypertension and diabetes mellitus. For patients with advanced CKD, the median age was 61 (IQR 51-69) years, eGFR 33 (30 -39) mL/min/1.73 m^2^, serum bicarbonate 22 (IQR 20 – 24), hemoglobin 12.9 (IQR 11.5 – 14.0) g/dl, serum transferrin 2.44 (IQR 2.23 – 2.73) g/L, serum uric acid 0.43 (IQR 0.37 – 0.53) and serum potassium 4.4 (IQR 3.9 – 4.8) mmol/L. The prevalence of metabolic acidosis was 62.4 %, anemia 46.4 %, gout 30.9 %, low transferrin levels 16.6 % and hyperkalemia 8.8 % among those with advanced CKD, while the prevalence of metabolic acidosis and anemia was 46.6 % and 25.9 % respectively in those with early CKD. Variables with higher odds for advanced CKD after multivariable logistic regression analysis were hypertension (OR 3.3, 95 % CI 1.2 - 9.2, P = 0.020), diabetes mellitus (OR 1.8, 95 % CI 1.1 - 3.3, P = 0.024), severe proteinuria (OR 3.5, 95 % CI 1.9 - 6.5, P = 0.001), angina (OR 2.5, 95 % CI 1.2 - 5.1, P = 0.008), anaemia (OR 2.9, 95% CI 1.7 - 4.9, P= 0.001), hyperuricemia (OR 2.4, 95 % CI 1.4 - 4.1, P = 0.001), and metabolic acidosis (OR 2.0, 95% CI 1.2 - 3.1, P= 0.005). Other associations with advanced CKD were widow/widower (OR 3.2, 95 % CI 1.4 - 7.4, P = 0.006), low transferrin (OR 2.4, 95% CI 1.1 - 5.1, P= 0.028), hyperkalemia (OR 5.4, 95% CI 1.2 - 24.1, P= 0.029), allopurinol (OR 2.4, 95 % CI 1.4 - 4.3, P = 0.005) and doxazosin (OR 1.9, 95% CI 1.2 - 3.1, P = 0.006).

**Conclusion:** Hypertension and diabetes mellitus were strongly associated with advanced CKD, suggesting a need for primary and secondary population-based prevention measures. Metabolic acidosis, anaemia with low transferrin levels, hyperuricemia and hyperkalemia were highly prevalent in our patients, including those with early CKD, and they were strongly associated with advanced CKD, calling for the proactive role of clinicians and dietitians in supporting the needs of CKD patients in meeting their daily dietary requirements towards preventing and slowing the progression of CKD.

## Introduction

Chronic kidney disease (CKD), defined as decreased kidney function identified by glomerular filtration rate (GFR) of less than 60 mL/min per 1·73 m^2^, or markers of kidney damage, or both, of at least 3 months duration, regardless of the underlying cause (1), is a major public health issue worldwide and contributes immensely to the overall non-communicable disease (NCD) burden, with NCDs also contributing to the burden of CKD (2, 3). Recent systematic reviews have reported the prevalence of CKD to be 15.8% in Africa, similar to other continents, constituting a true public health need with major cost implications to healthcare systems (1, 4). Chronic kidney disease is usually asymptomatic until the more advanced stages and accurate prevalence data are lacking in most regions including sub-Saharan Africa (5). As the prevalence of CKD is increasing worldwide and consequently the demand for kidney replacement therapy (KRT), the incidence of cardiovascular events and death is also increasing (6-8). Diabetes mellitus and hypertension are the leading causes of CKD worldwide; and in sub-Saharan Africa, hypertension is the leading cause of CKD (9, 10).

In addition to the known risk factors for advanced CKD, which are age, male sex, black race, arterial hypertension and proteinuria, other modifiable factors including medications (traditional and herbal), hyperuricemia, hyperlipidemia, elevated phosphate levels, heart failure and anemia are common in CKD at later stages (11-13). Metabolic acidosis increases with worsening eGFR, with prevalence of around 40% among patients with CKD stage 4 and it is associated with rapid CKD progression (14-16). Anemia is common in CKD and is frequently associated with poor outcomes, including increased cardiovascular risks, hospitalization, decreased quality of life and increased risk of mortality (17-19). Hyperuricemia greatly contributes to the development of CKD and its progression despite the fact that there are no clear cut-off levels for uric acid that are associated with the risk for CKD. It appears that increasing uric acid levels increase the risk for CKD development by causing inflammation, endothelial cell injury and activation of the renin-angiotensin system (20-22). Hyperkalemia is also common in advanced CKD; its prevalence increases with decreasing eGFR and it is significantly associated with faster CKD progression (23, 24).

Individuals of black ethnicity due to their genetics, including the presence of APOL1 high risk genotypes, are at higher risk of death due to CKD as a result of social, economic and medical causes (25, 26). African ancestry has been associated with higher serum creatinine levels, lower eGFR estimates and more rapid CKD progression (27, 28). Thus, the aim of this study was to determine the demographic and clinical profile of black patients with CKD attending Charlotte Maxeke Johannesburg Academic Hospital (CMJAH) in Johannesburg, South Africa.

## Methods

### Study design, population and settings

This was a cross sectional study to evaluate the demographic and clinical profile of black patients with CKD attending the kidney outpatient department (KOPD) clinic at CMJAH between September 2019 to March 2020. Inclusion criteria included patients who were >18 years of age, CKD stages 1 – 4, who had controlled hypertension (blood pressure < 140/90 mm Hg) and diabetes mellitus (HbA1C < 7%), attending the KOPD clinic for at least 6 months and were able to provide informed consent. Patients who had active infections, active malignancies, autoimmune diseases and who were not black were excluded. Johannesburg is the largest city in South Africa and among the largest 50 urban agglomerations in the world. Johannesburg had an estimated population of 5.9 million in 2021 with th<u>e most common racial groups being</u> black African (76.4%), colored (5.6%), White (12.3%) and Indian/Asian (4.9%). The illiterate account for 7% of the population and 3.4% has only primary school education, and 29% of residents live in informal dwellings. Due to increased urbanization, there has been increased use of westernized dietary patterns often comprising of fast and deep-fried foods, processed meats, saturated fats, sugar sweetened beverages, alcoholic beverages plus other highly processed foods, unlike in rural areas where the traditional dietary patterns are predominant (29).

### Data collection and laboratory procedures

Demographic and clinical data including age, gender, weight, height, glycemic status, history of smoking, etiology of CKD and medications were extracted from the ongoing continuous KOPD clinic records and face to face interviews, and were filled in a questionnaire. Systolic and diastolic blood pressure was measured 3 times, the average of the second and third measurements was used. Body mass index (BMI) was calculated using the National Health Services (NHS-UK) BMI calculator(30). Measurements of urinary protein creatinine ratio (uPCR), serum creatinine, electrolytes, HbA1C, WBC, haemoglobin level, platelets, calcium, phosphate, transferrin and HDL cholesterol were done as standard of care at the time of recruitment during a clinic visit. Serum creatinine was measured using the isotope dilution mass spectrometry (IDMS) traceable enzymatic assay and estimated glomerular filtration rate (eGFR) was calculated using the Chronic Kidney Disease Epidemiology Collaboration (CKD-EPI) equation without using the African American correction factor (31). Patients with eGFR < 45 ml/min/1.72m^2^ were considered to have advanced CKD, while those patients with eGFR ≥ 45 ml/min/1.72m^2^ early CKD.

### Data management and analysis

Data collected was entered into a REDCap (Research Electronic Data Capture) (Vanderbilt University, 2014) database and analyzed using STATA version 17 (College Station, Texas, USA). Descriptive statistics were used to summarize demographic and clinical characteristics; continuous variables have been reported as medians with interquartile ranges and Wilcoxon rank-sum test was used for the non-normally distributed variables. Discrete variables have been reported as frequencies and proportions, Pearson’s chi-square test were used to test for association between two variables. Odd ratios were used to estimate the strengths of association between variables and advanced CKD, univariate and multivariate logistic regression models have been used to estimate the association of variables and advanced CKD. Variables with p-value less than 0.2 on univariate logistic regression models were then fitted into the multivariate logistic regression models with the addition of age and sex as adjusting variables; variables with a p-value of less than 0.05 were considered to have significant strength of association.

### Ethical issues

Ethical approval was obtained from the Human Research Ethics Committee of the University of Witwatersrand, Johannesburg (ethics clearance certificate No. M190553). Informed consent was obtained from each of the participants before embarking on data collection.

## Results

### Demographic and clinical characteristics of the study population

Of the 476 black patients with CKD stage 1-4 who attended the CMJAH KOPD clinic during the study period, 164 patients were excluded from the study including 110 patients who had uncontrolled hypertension, 35 patients who had uncontrolled diabetes mellitus, 11 patients had autoimmune diseases, 6 patients had active infections and 2 patients had active malignancies. A total of 312 CKD black patients were enrolled into this study, of whom 162 (51.9 %) were male, 292 (93.6%) were hypertensive, 103 (33.0%) were diabetic and 164 (52.6 %) were married. The median age was 61 (IQR 51-69) years for advanced CKD and 53 (IQR 41 -62) years for early CKD; the median eGFR was 33 (30 -39) mL/min/1.73 m^2^ for advanced CKD and 60 (IQR 51 – 75) mL/min/1.73 m^2^ for early CKD; the median urine protein creatinine ratio (uPCR) was 0.029 (IQR 0.015 -0.67) g/mmol for advanced CKD and 0.016 (IQR 0.008 – 0.034) g/mmol for early CKD. The median serum bicarbonate was 22 (IQR 20 – 24) mmol/L for advanced CKD and 23 (IQR 21 -25) mmol/L for early CKD; the median hemoglobin (Hb) was 12.9 (IQR 11.5 – 14.0) g/dl for advanced CKD and 13.8 (IQR 12.4 – 15.7) g/dl for early CKD; the median serum transferrin was 2.44 (IQR 2.23 – 2.73) g/L for advanced CKD and 2.62 (IQR 2.37 – 2.89) g/L for early CKD. The median serum uric acid was 0.43 (IQR 0.37 – 0.53) mmol/L for advanced CKD and 0.36 (IQR 0.30 – 0.46) mmol/L for early CKD (Table 1).

**Table 1:** Demographic characteristics and clinical profile of 312 CKD patients by eGFR.

| Characteristic | eGFR < 45 ml/min/1.72m<sup>2</sup><br>(n=181) | eGFR ≥ 45 ml/min/1.72m<sup>2</sup><br>(n= 131) | P-value |
| --- | --- | --- | --- |
|  | Proportion (%) or Median (IQR) | Proportion (%) or Median (IQR) |  |
| Age (years) | 61 (51-69) | 53 (41 -62) | **0.001** |
| **Sex** |  |  |  |
| Male | 96 (53.0%) | 66 (50.4%) | 0.643 |
| Female | 85(47.0%) | 65 (49.6%) |  |
| **Marital status** |  |  |  |
| Single | 43(23.8%) | 48(36.6%) | **0.026** |
| Married | 98 (54.1%) | 66 (50.4%) |  |
| Widow/Widower | 30(16.6%) | 10 (7.6%) |  |
| Separated/Divorced | 10(5.5%) | 7 (5.4%) |  |
| **Highest level of education** |  |  |  |
| No formal education | 24(13.3%) | 14 (10.7%) | 0.505 |
| Primary | 41(22.7%) | 30 (22.9%) |  |
| Secondary | 56(30.9%) | 43 (32.8%) |  |
| Tertiary | 60(33.1%) | 44 (33.6%) |  |
| **Occupation** |  |  |  |
| Unemployed | 26(14.4%) | 19(14.5%) | 0.523 |
| Domestic workers | 37(20.4%) | 25(19.1%) |  |
| Self employed | 43(23.8%) | 27(20.6%) |  |
| Public / Private servant | 57(31.5%) | 52(39.7%) |  |
| Retired | 18(9.9%) | 8(6.1%) |  |
| BMI (kg/m<sup>2</sup>) | 30.2 (26.0 -34.5) | 30.2 (26.6 - 34.7) | 0.731 |
| SBP (mmHg) | 140 (130 – 140) | 140 (128 -140) | 0.339 |
| DBP(mmHg) | 82 (72 – 90) | 83 (74 -90) | 0.630 |
| uPCR (g/mmol) | 0.029 (0.015 -0.67) | 0.016 (0.008 – 0.034) | **0.001** |
| Creatinine (umol/L) | 163 (141-190) | 109 (88 -122) | **0.001** |
| eGFR (ml/min/1.72m<sup>2</sup>) | 33 (30 -39) | 60 (51 – 75) | **0.001** |
| FBG (mmol/L) | 4.5 (4.2 – 5.2) | 4.4 (4.2 – 4.9) | 0.146 |
| HbA1C (%) | 7.0 (6.6 – 7.0) | 7.0 (7.0 – 7.0) | 0.222 |
| Haemoglobin (g/dl) | 12.9 (11.5 – 14.0) | 13.8 (12.4 – 15.7) | **0.001** |
| Transferrin (g/L) | 2.44 (2.23 – 2.73) | 2.62 (2.37 – 2.89) | **0.001** |
| WBC (x 10<sup>9</sup> cells/L) | 6.4 (5.13 – 7.71) | 6.03 (4.81 – 7.73) | 0.420 |
| Platelets (x 10<sup>9</sup> cells/L) | 256 (213 -320) | 271 (217 – 325) | 0.378 |
| Uric acid (mmol/L) | 0.43 (0.37 – 0.53) | 0.36 (0.30 – 0.46) | **0.001** |
| HDL cholesterol (mmol /L) | 1.23 (0.98 -1.48) | 1.22 (1.02 – 1.52) | 0.528 |
| Calcium (mmol /L) | 2.31 (2.23 – 2.40) | 2.33 (2.26 – 2.42) | 0.054 |
| Phosphate (mmol /L) | 1.1 (0.94 – 1.24) | 1.02 (0.85 – 1.15) | **0.005** |
| Sodium (mmol/L) | 141 (138 – 143) | 141 (139 – 143) | 0.965 |
| Potassium (mmol/L) | 4.4 (3.9 – 4.8) | 4.1 (3.9 – 4.4) | **0.001** |
| Bicarbonate (mmol/L) | 22 (20 – 24) | 23 (21 -25) | **0.013** |

IQR, interquartile range; uPCR, urine protein creatinine ratio; DBP, diastolic blood pressure; eGFR, estimated glomerular filtration rate; FBG, fasting blood sugar; HbA1C, glycosylated hemoglobin A1C; WBC, white blood cells; HDL, high density lipoprotein; SBP, systolic blood pressure.

### Clinical profile of CKD patients

Of the 312 CKD black patients, 58% patients had advanced CKD (CKD stage 3b or 4), of whom 57 (31.5 %) patients presented with severely increased proteinuria as compared to 23 (17.4 %) patients with early CKD. Metabolic acidosis was present in 113 (62.4 %) of those who had advanced CKD and in 61 (46.6 %) patients with early CKD. Anaemia was present in 84 (46.4 %) patients with advanced CKD including 30 (16.6 %) patients who had low transferrin levels, while 34 (25.9 %) patients with early CKD had anemia. Among patients with advanced CKD, hyperuricemia was found in 56 (30.9 %) patients and 16 (8.8 %) patients had hyperkalemia. Among patients with advanced CKD, majority (96.7 %) patients were diagnosed with hypertension, 70 (38.7 %) patients had diabetes mellitus and 69 (38.1%) had both hypertension and diabetes mellitus as compared to 117 (89.3 %) patients who had hypertension, 33 (25.2 %) patients who had diabetes mellitus and 32 (24.4%) had both hypertension and diabetes mellitus among those with early CKD. Angina was reported in 41 (22.7%) patients with advanced CKD and 15 (11.5 %) patients in early CKD. Most (56.9 %) patients with advanced CKD were using more than 5 medications as compared to 41.2 % patents with early CKD who were using 3 – 4 medications for their blood pressure control. Majority (84.0 %) of the patients with advanced CKD and 76.3 % patients in early CKD were on calcium channel blockers (CCBs), while 18.8 % patients with advanced CKD and 21.4 % of those with early CKD were using angiotensin converting enzyme inhibitors (ACEIs) or angiotensin receptors blockers (ARBs). For patients with advanced CKD, most (57.5 %) were on diuretics followed by 51.4 % on doxazosin; 29.3 % of patients were on insulin and 16 (8.8 %) on oral hypoglycemic agents (Table 2).

**Table 2:** Clinical profile of 312 CKD patients by eGFR.

| Parameter | eGFR < 45<br>ml/min/1.72m<sup>2</sup> (n=181)<br>Proportion (%) | eGFR ≥ 45<br>ml/min/1.72m<sup>2</sup> (n= 131)<br>Proportion (%) | P-value |
| --- | --- | --- | --- |
| **eGFR (ml/min/1.72m<sup>2</sup>)** |  |  |  |
| Stage 1 (> 90) | 0 (0.0 %) | 4 (3.1 %) |  |
| Stage 2 (60 – 89) | 0 (0.0 %) | 62 (47.3 %) |  |
| Stage 3a (45 -59) | 0 (0.0 %) | 65 (49.6 %) |  |
| Stage 3b (30-44) | 144 (79.6 %) | 0 (0.0 %) |  |
| Stage 4 (16- 29) | 37 (20.4 %) | 0 (0.0 %) | **0.001** |
| **Diagnoses encountered** |  |  |  |
| Hypertension | 175 (96.7 %) | 117 (89.3 %) | **0.009** |
| Diabetes mellitus | 70 (38.7 %) | 33 (25.2 %) | **0.012** |
| Hypertension & Diabetes mellitus | 69 (38.1%) | 32 (24.4%) | **0.011** |
| Adult polycystic kidney disease | 5 (2.8 %) | 7 (5.3 %) | 0.242 |
| Reflux nephropathy | 0 (0.0 %) | 5 (3.8 %) | **0.008** |
| Obstructive uropathy | 1 (0.6 %) | 0 (0.0 %) | 0.394 |
| Unknown | 6 (3.3 %) | 5 (3.8 %) | 0.812 |
| **Current smoking** |  |  |  |
| Yes | 13 (7.2%) | 10 (7.6%) |  |
| No | 168 (92.8%) | 121 (92.4%) | 0.880 |
| **Current Alcohol** |  |  |  |
| Yes | 14 (7.7%) | 18 (13.7%) | 0.084 |
| No | 169 (92.3%) | 113 (86.3%) |  |
| **Cardiovascular Diseases** |  |  |  |
| None | 120 (66.3%) | 108 (82.4 %) | 0.017 |
| Angina | 41 (22.7%) | 15 (11.5 %) |  |
| Myocardial infarction | 8 (4.4%) | 4 (3.0 %) |  |
| Heart failure | 6 (3.3%) | 2 (1.5 %) |  |
| Stroke | 6 (3.3%) | 1 (0.8 %) |  |
| Transient ischemic attack | 0 (0.0%) | 1 (0.8 %) |  |
| **Medications** |  |  |  |
| None | 2 (1.1 %) | 8 (6.1 %) | 0.004 |
| Diuretics | 104 (57.5 %) | 58 (44.3 %) | 0.021 |
| ACEIs / ARBs | 34 (18.8 %) | 28 (21.4 %) | 0.572 |
| Aldactone | 6 (3.3 %) | 5 (3.8 %) | 0.812 |
| CCBs | 152 (84.0 %) | 100 (76.3 %) | 0.092 |
| Statins | 100 (55.3 %) | 60 (45.8 %) | 0.099 |
| Oral hypoglycemics | 16 (8.8 %) | 21(16.0 %) | 0.053 |
| Insulin | 53 (29.3 %) | 19 (14.5 %) | 0.002 |
| Allopurinol | 22 (12.1 %) | 13 (9.9 %) | 0.538 |
| Junior ASA | 90 (49.7 %) | 57 (43.5 %) | 0.278 |
| Beta blockers | 41 (22.7 %) | 30 (22.9 %) | 0.959 |
| Aldomet | 12 (6.6 %) | 7 (5.3 %) | 0.639 |
| Hydralazine | 4 (2.2 %) | 3 (2.3 %) | 0.962 |
| Doxazosin | 93 (51.4 %) | 48 (36.6 %) | 0.010 |
| Others | 68 (37.6 %) | 38 (29.0 %) | 0.115 |
| **Number of medications per patient** |  |  |  |
| 0 | 2 (1.1 %) | 8 (6.1 %) | 0.001 |
| 1 - 2 | 20 (11.1 %) | 20 (15.3 %) |  |
| 3 – 4 | 56 (30.9 %) | 55 (41.2 %) |  |
| ≥ 5 | 103 (56.9 %) | 48 (36.6 %) |  |
| **uPCR (g/mmol)** |  |  |  |
| Normal to mildly increased (<0.015) | 47 (26.0 %) | 63 (48.2 %) | 0.001 |
| Moderately increased (0.015- 0.05) | 77 (42.5 %) | 45 (34.4 %) |  |
| Severely increased (> 0.050) | 57 (31.5 %) | 23 (17.4 %) |  |
| **Haemoglobin (g/dl)** |  |  |  |
| Normal (> 12.5) | 97 (53.6 %) | 97 (74.1 %) | 0.001 |
| Anemia (<12.5) | 84 (46.4 %) | 34 (25.9 %) |  |
| **Transferrin g/dl)** |  |  |  |
| Low (< 2.0) | 30 (16.6 %) | 10 (7.6 %) | 0.052 |
| Normal (2.0-3.60) | 148 (81.8 %) | 117 (89.3 %) |  |
| High (>3.60) | 3 (1.7 %) | 4 (3.1 %) |  |
| **Uric acid (mmol/l)** |  |  |  |
| Low (< 0.16 or 0.21) | 4 (2.2 %) | 15 (11.5 %) | 0.001 |
| Normal (0.16/0.21 – 0.36/0.43) | 121 (66.9 %) | 98 (74.8 %) |  |
| High (> 0.36 or 0.43) | 56 (30.9 %) | 18 (13.7 %) |  |
| **Potassium (mmol/l)** |  |  |  |
| Low (< 3.5) | 13 (7.2 %) | 15 (11.5 %) |  |
| Normal (3.5 – 5.1) | 152 (84.0 %) | 114 (87.0 %) | **0.013** |
| High (> 5.1) | 16 (8.8 %) | 2 (1.5 %) |  |
| **Bicarbonate (mmol/l)** |  |  | **0.018** |
| Low (< 23) | 113 (62.4 %) | 61 (46.6 %) |  |
| Normal (23 – 29) | 67 (37.0 %) | 68 (51.9 %) |  |
| High (> 29) | 1 (0.6 %) | 2 (1.5 %) |  |

uPCR, urine protein creatinine ratio; eGFR, estimated glomerular filtration rate; CCBs, calcium channel blockers; ACEIs, angiotensin converting enzyme inhibitors; ARBs, Aldosterone receptors blockers.

### Factors associated with advanced CKD

We divided the patients into two subgroups according to their CKD stages [early CKD (eGFR ≥ 45 ml/min/1.72m^2^) vs. advanced CKD (eGFR < 45 ml/min/1.72m^2^)]. A total of 24 potential variables were identified after performing univariate logistic regression analyses. Backward elimination reduced this to 15 parameters; the factors associated with advanced CKD after adjusting for age and sex on multivariate logistic regression analysis included: hypertension (OR 3.3, 95 % CI 1.2 - 9.2, P = 0.020), diabetes mellitus (OR 1.8, 95 % CI 1.1 - 3.3, P = 0.024), angina (OR 2.5, 95 % CI 1.2 - 5.1, P = 0.008), severe proteinuria (OR 3.5, 95 % CI 1.9 - 6.5, P = 0.001), moderate proteinuria (OR 2.5, 95% CI 1.5 - 4.3, P=0.001), hyperuricemia (OR 2.4, 95 % CI 1.4 - 4.1, P = 0.001), anaemia (OR 2.9, 95% CI 1.7 - 4.9, P= 0.001), metabolic acidosis(OR 2.0, 95% CI 1.2 - 3.1, P= 0.005), allopurinol (OR 2.4, 95 % CI 1.4 - 4.3, P = 0.005) and doxazosin (OR 1.9, 95% CI 1.2 - 3.1, P = 0.006), low transferrin (OR 2.4, 95% CI 1.1 - 5.1, P= 0.028), hyperkalemia (OR 5.4, 95% CI 1.2 - 24.1, P= 0.029) and, widow/widower (OR 3.2, 95 % CI 1.4 - 7.4, P = 0.006) (Tables 3).

**Table 3:** Factors associated with advanced CKD.

| Characteristic | Number of patients | UNIVARIATE |  | MULTIVARIATE |  |
| --- | --- | --- | --- | --- | --- |
|  |  | OR (95% CI) | P. value | OR (95% CI) | P. value |
| **Age** | 312 | 1.1(1.0-1.2) | 0.001 |  |  |
| **Marital status** |  |  |  |  |  |
| Single | 91 | 1(Reference) |  | 1(Reference) |  |
| Married | 164 | 1.7 (0.9-2.8) | 0.055 | 1.5 (0.9-3.0) | 0.137 |
| Widow/Widower | 40 | 3.3 (1.5-7.7) | 0.004 | 3.2 (1.4-7.4) | **0.006** |
| Separated/Divorced | 17 | 1.6 (0.6-4.6) | 0.038 | 1.4 (0.5-4.3) | 0.466 |
| **Hypertension** |  |  |  |  |  |
| NO | 20 | 1(Reference) |  | 1(Reference) |  |
| YES | 292 | 3.5 (1.3 – 9.3) | 0.013 | 3.3 (1.2-9.2) | **0.020** |
| **Diabetes mellitus** |  |  |  |  |  |
| NO | 209 | 1(Reference) |  | 1(Reference) |  |
| YES | 103 | 1.9 (1.1 – 3.1) | 0.013 | 1.8 (1.1-3.0) | **0.024** |
| **Current Alcohol** |  |  |  |  |  |
| No | 280 | 1(Reference) |  | 1(Reference) |  |
| Yes | 32 | 0.5 (0.3-1.1) | 0.088 | 0.5 (0.2-1.0) | 0.059 |
| **Cardiovascular Disease:** |  |  |  |  |  |
| None | 228 | 1(Reference) |  | 1(Reference) |  |
| Angina | 56 | 2.7 (1.4-5.3) | 0.004 | 2.5 (1.2-5.1) | **0.008** |
| Myocardial infarction | 12 | 1.8 (0.5-6.1) | 0.348 | 1.4 (0.4-5.0) | 0.578 |
| Heart failure | 8 | 2.7 (0.5-13.7) | 0.230 | 2.8 (0.5-14.5) | 0.219 |
| Stroke | 8 | 5.4 (0.6-45.6) | 0.121 | 4.6 (0.5-39.2) | 0.167 |
| **Medications** |  |  |  |  |  |
| No | 10 | 1 (Reference) |  | 1(Reference) |  |
| Yes | 302 | 11.7 (1.4-94.8) | 0.021 | 0.1 (0.0-0.9) | **0.042** |
| **Diuretics** |  |  |  |  |  |
| No | 150 | 1(Reference) |  | 1(Reference) |  |
| Yes | 162 | 1.7 (1.1-2.7) | 0.022 | 1.6 (1.0-2.5) | 0.053 |
| **CCB** |  |  |  |  |  |
| No | 60 | 1(Reference) |  | 1(Reference) |  |
| Yes | 252 | 1.6 (0.9-2.9) | 0.093 | 1.6 (0.9-2.8) | 0.127 |
| **Statins** |  |  |  |  |  |
| No | 152 | 1(Reference) |  | 1(Reference) |  |
| Yes | 160 | 1.5 (0.9-2.3) | 0.100 | 1.4 (0.9-2.2) | 0.171 |
| **Oral hypoglycemics** |  |  |  |  |  |
| No | 275 | 1(Reference) |  | 1(Reference) |  |
| Yes | 37 | 0.5 (0.3-1.0) | 0.056 | 0.5 (0.2-1.0) | **0.048** |
| **Allopurinol** |  |  |  |  |  |
| No | 277 | 1(Reference) |  | 1(Reference) |  |
| Yes | 35 | 2.5 (1.4 – 4.4) | 0.003 | 2.4 (1.3-4.3) | **0.005** |
| **Nitrates** |  |  |  |  |  |
| No | 306 | 1(Reference) |  | 1(Reference) |  |
| Yes | 6 | 1.9 (1.1-3.3) | 0.024 | 1.7 (1.0-3.0) | 0.059 |
| **Doxazosin** |  |  |  |  |  |
| No | 171 | 1(Reference) |  | 1(Reference) |  |
| Yes | 141 | 1.8 (1.2-2.9) | 0.010 | 1.9 (1.2-3.1) | **0.006** |
| **Others** |  |  |  |  |  |
| No | 206 | 1(Reference) |  | 1(Reference) |  |
| Yes | 106 | 1.5 (0.9-2.4) | 0.116 | 1.5 (0.9-2.5) | 0.100 |
| **Proteinuria (g/mmol)** |  |  |  |  |  |
| Normal to mild (<0.015) | 110 | 1(Reference) |  | 1(Reference) |  |
| Moderately (0.015- 0.05) | 122 | 2.3 (1.4 – 3.9) | 0.002 | 2.5 (1.5- 4.3) | **0.001** |
| Severely (> 0.050) | 80 | 3.9 (2.0 – 7.2) | 0.000 | 3.5 (1.9 - 6.5) | **0.001** |
| **Hyperuricemia** |  |  |  |  |  |
| No | 216 | 1(Reference) |  | 1(Reference) |  |
| YES | 96 | 2.4 (1.4 - 4.0) | 0.001 | 2.4 (1.4 - 4.1) | **0.001** |
| **Anaemia** |  |  |  |  |  |
| No | 194 | 1(Reference) |  | 1(Reference) |  |
| YES | 118 | 2.5 (1.5 - 4.0) | 0.001 | 2.9 (1.7 - 4.9) | **0.001** |
| **Transferrin g/dl)** |  |  |  |  |  |
| Low (< 2.0) | 40 | 1(Reference) |  | 1(Reference) |  |
| Normal (2.0-3.60) | 265 | 2.4 (1.1 - 5.1) | 0.025 | 2.4 (1.1- 5.1) | **0.028** |
| High (>3.60) | 7 | 0.6 (0.1 - 2.7) | 0.499 | 0.5 (0.1- 2.7) | 0.491 |
| **WBC (x 10<sup>9</sup> cells/l)** |  |  |  |  |  |
| Low (> 4) | 23 | 1(Reference) |  | 1(Reference) |  |
| Normal (4-11) | 276 | 0.5 (0.2 - 1.3) | 0.051 | 1.8 (0.7 - 4.2) | 0.202 |
| High (>11) | 13 | 1.1 (0.4 - 3.5) | 0.859 | 1.8 (0.4 - 7.5) | 0.418 |
| **Calcium (mmol /l)** |  |  |  |  |  |
| Low (< 2.15) | 25 | 1(Reference) |  | 1(Reference) |  |
| Normal (2.15 -2.45) | 244 | 2.2 (0.9 - 5.8) | 0.098 | 2.1 (0.8 - 5.8) | 0.125 |
| High (>2.45) | 43 | 0.6 (0.3 - 1.1) | 0.081 | 0.6 (0.3 - 1.1) | 0.081 |
| **Potassium (mmol/l)** |  |  |  |  |  |
| Low (< 3.5) | 28 | 1(Reference) |  | 1(Reference) |  |
| Normal (3.5 – 5.1) | 266 | 0.7 (0.3 - 1.4) | 0.280 | 0.6 (0.3 - 1.4) | 0.261 |
| High (> 5.1) | 18 | 6.0 (1.4 - 26.6) | 0.018 | 5.4 (1.2 -24.1) | **0.029** |
| **Bicarbonate (mmol/l)** |  |  |  |  |  |
| Low (< 23) | 174 | 1(Reference) |  | 1(Reference) |  |
| Normal (23 – 29) | 135 | 1.9 (1.2 - 3.0) | 0.007 | 2.0 (1.2 - 3.1) | **0.005** |
| High (> 29) | 3 | 0.5 (0.1 - 5.7) | 0.583 | 0.4 (0.0 - 5.1) | 0.495 |

P < 0.05 was used to identify potential variables, those with P <0.2 in univariate were included into the multivariate analysis while adjusting for age and sex.

## Discussion

This study evaluated the demographic and clinical profile of black patients with CKD attending the CMJAH kidney outpatient clinic in Johannesburg, South Africa. There were 42% with early CKD and 58% with advanced CKD, of whom 93.6% had hypertension and 33.0 % diabetes mellitus, and 32.3% had both hypertension and diabetes. The majority (96.7 %) of patients with advanced CKD had hypertension, similar to findings in other studies from SSA (9, 10, 32, 33). Approximately 51.4 % of patients with advanced CKD were using doxazosin for treatment of hypertension with a 1.9 times increased association with advanced CKD, as also been reported in other studies (34, 35). Peripherally acting α-blockers like doxazosin are commonly used in the management of hypertension in CKD, mainly due to their pharmacokinetic profile that is undisturbed by worsening kidney function and their role in blood sugar control (36). Approximately 38.7 % patients with advanced CKD had T2DM; T2DM had 1.8 increased risk for advanced CKD, similar to other studies conducted among black patients in South Africa, Tanzania and Ethiopia (1, 9, 33, 37). In patients with advanced CKD, approximately 29.3 % of the patients were on insulin and 8.8 % were on oral hypoglycemics for treatment of their T2DM. Oral hypoglycemics were associated with 0.5 times higher risk for advanced CKD, similar to other studies (38, 39).

The prevalence of metabolic acidosis was 62.4 % in advanced CKD and 46.6 % in early CKD; this prevalence is higher than the 33% - 40% among patients with CKD stage 3 – 4 from other continents (14, 40, 41). The possible explanation could be firstly, the more rapid CKD progression which has been shown to occur in black patients even early in their CKD stages (16, 28) and secondly, diet where replacement of traditional diets with contemporary/ western foods which contain mainly animal proteins, less vegetables and low intake of fruits might increase CKD patients’ dietary acid load (29, 42-45). Patients with low serum bicarbonate levels were 2-fold more likely to have advanced CKD, as reported also in other studies (14, 15). The prevalence of anaemia was 46.4 % in advanced CKD, including 16.6 % who had low transferrin levels, while 25.9 % of the patients with early CKD had anemia. Advanced CKD was 2.9 times more prevalent if a patient had anemia and 2.4 times more prevalent if a patient had low transferrin, similar to other studies (17-19). The prevalence of hyperuricemia was 30.9 % in advanced CKD; advanced CKD had a 2.4-fold higher OR if a patient had hyperuricemia and 2.4-fold higher if a patient was on allopurinol; similar findings have been reported in other studies (11) (20-22). Approximately 8.8 % patients with advanced CKD were found to have hyperkalemia; hyperkalemia had a 5.4-fold increased association with advanced CKD; similar findings have been reported in other studies (23, 24).

Studies have shown that the incidence of cardiovascular events increases with worsening kidney function (46, 47); 22.7% patients with advanced CKD reported angina with a 2.5 times increased risk in advanced CKD, similar to other studies (46, 48). Also 31.5 % patients with advanced CKD presented with severely increased proteinuria; advanced CKD was 3.5 times higher if a patient had severe proteinuria, as also reported in other studies (3, 13, 37, 49). Furthermore, few (18.8 %) patients with advanced CKD and 21.4 % of those with early CKD were using ACEIs or ARBs; similar findings have been reported from other studies on the underutilization of ACEIs/ARBs when they were clinically indicated or discontinuation of RAAS inhibitors by clinicians during the course of CKD possibly due to their associated side effects (50-52). Angiotensin converting enzyme inhibitors (ACEIs) or angiotensin receptors blockers (ARBs) had no significant association with advanced CKD, possibly due to the small numbers on these agents, unlike findings from other studies that did demonstrate that the use of ACEIs/ARBs had beneficial effects for kidney events and cardiovascular outcomes compared to other antihypertensive medications in patients with CKD (52-54). The possible explanation could be the fact that the majority (84.0 %) of the study patients with advanced CKD were using calcium channel blockers. Studies have demonstrated that calcium channel blockers have similar kidney and cardiovascular protective effects when compared to RAAS blockers in patients with CKD (55, 56).

## Conclusion

Hypertension and diabetes mellitus were strongly associated with advanced CKD, suggesting a need for primary and secondary population-based prevention measures. Metabolic acidosis, anemia with low transferrin levels, hyperuricemia and hyperkalemia were highly prevalent in our patients, including those with early CKD, and they were strongly associated with advanced CKD. This calls for the proactive role of clinicians and dietitians in supporting the needs of CKD patients in meeting their daily dietary requirements towards preventing and slowing the progression of CKD. Further studies on the role of diet including plant-based proteins, vegetables and fruits in preventing and slowing CKD progression and other metabolic complications of CKD among black patients are warranted.

## Data Availability

All relevant data are within the manuscript and its Supporting Information files.

## Abbreviations

CKD: chronic kidney diseases
CMJAH: Charlotte Maxeke Johannesburg Academic Hospital
CKD-EPI: chronic kidney disease epidemiology collaboration
ESKD: end stage kidney disease
T2DM: Type 2 Diabetes Mellitus
eGFR: estimated glomerular filtration rate
IDMS: isotope dilution mass spectroscopy
uPCR: urine protein creatinine ratio
CCBs: calcium channel blockers
ACEIs: angiotensin converting enzyme inhibitors
ARBs: Aldosterone receptors blockers
KDIGO: kidney disease improving global outcomes
NCDs: non-communicable diseases
SSA: sub Saharan Africa

## Acknowledgements

Special thanks to all patients with CKD attending at the Charlotte Maxeke Johannesburg Academic Hospital (CMJAH) kidney outpatient clinic for their willingness to participate in this study and to the staff at the Charlotte Maxeke Johannesburg Academic Hospital (CMJAH) kidney outpatient clinic and laboratory for their continued care, proper keeping of patient data and support which made this study possible. I thank the International Society of Nephrology (ISN), the University of the Witwatersrand, the University of Dodoma and Shinei industries (Aichi, Japan) Co. Ltd, for support, and providing a good and conducive environment for my nephrology fellowship training.

## Authors’ contributions

Design of the work: AJM, SN, RD &

GP. Data collection: AJM, SN, RD, GP & TD.

Data processing & analysis: AJM, SN, RD, CD & DB.

Manuscript writing: AJM, SN, RD, GP, CD, TD & DB

All authors have read and approved the final manuscript.

